# The impact of vaccination on the spread patterns of the COVID epidemic

**DOI:** 10.1101/2021.04.29.21256322

**Authors:** D. Below, F. Mairanowski

## Abstract

A modified model of the epidemic under conditions of mass vaccination was developed. A comparison of the model results with statistical observations in Israel shows good agreement.

Model calculations are performed on the efficacy of limiting the development of an epidemic by both lockdown and vaccination. Mass vaccination of the population is the most radical method of limiting the growth of the epidemic. The introduction of a lockdown cannot completely prevent the development of an epidemic. The likelihood of the emergence of new strains of the virus is assessed. Without vaccination, the probability of more than two new virus strains per year affecting the epidemic growth process is found to be about 60%. A controlled calculation was made of the effect of the timing of changes in lockdown conditions during the vaccination period on the development of the epidemic. It was particularly shown that the cancellation of the lockdown together with the start of vaccination did not reduce the maximum number of new infections. A controlling calculation was made of the effects of gradually cancelling lockdown. On the basis of these calculations, it is possible to assess the development of the epidemic in different variants of partial lockdown cancellation.

Three dimensionless complexes, made up of the intensities of transmission, vaccination and lockdown restrictions, are found to determine the epidemic’s development.

The intensity of the coronavirus epidemic depends on climatic characteristics, in particular air temperature and the UV index. A relationship is given to estimate the influence of these factors on infection growth.

The way forward for further development of the model is outlined. The immediate goal of modifying the model is to use it for each age group in the population and to find out the links between vaccination rates and the psychological state of the population, i.e. people’s readiness for mass vaccination.

## Introduction

The numerous models for calculating epidemic spread can be divided into two broad groups, which differ fundamentally from each other. One group of models is based on differential equations describing changes with time in the number of infected individuals depending on the susceptible part of the population. They vary in their level of detail; however, they all require numerical methods for estimating the spread of the epidemic. Greater detailed elaboration of the models requires the introduction of an increasing number of empirical coefficients, which may not always adopt stable values for various specific cases. A detailed description of such models can be found in [1]. Another, stochastic group of models describes the probability of transmission through contact, the number of which is calculated by means of special sophisticated algorithms. These models, however, also require the introduction of a large number of empirical constants [2].

It is tempting to develop models that, by simplifying the modelling approach, allow analytical solutions to be obtained while taking into account the most important properties of a real spreading epidemic, enabling administrations to rapidly analyse changes in the epidemiological situation depending on the management decisions taken.

An example of such a model is the propagation analysis model [3]. The model was developed with the assumption that it can be used mainly at the initial stage of an epidemic, but further studies have shown its high accuracy also in the case of a mature epidemic.

## Methodology

The initial system of equations, which was the basis for the proposed model, is of the form [3]:

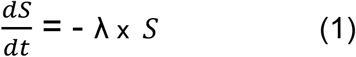

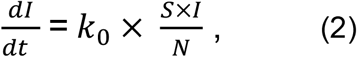

Where:

I - number of infected persons at a given time,

*k*_0_ - coronavirus infection rate (1/day)

N - total population of the area under consideration,

S - number of susceptible part of the population potentially capable of becoming infected due to contact with infected individuals.

λ - intensity factor of decrease in contacts of infected patients with persons who potentially can get infected by means of quarantine and other preventive measures.

In contrast to previously developed models of this type, in our model there is no inverse relationship between the values of I and S, i.e. under lockdown conditions and provided that I ≪ S (usually I ≤ 0.2 S), we can assume that the number of potentially susceptible persons does not vary with the number of infections and depends only on the severity of the lockdown:

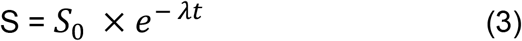

Where:

*S*_0_−is the maximum number of potentially infectious persons.

It has been shown in [4], [5], and [6] that switching from the absolute number of infected to their relative percentage per inhabitant yields universal dependencies for populated areas with substantially different populations.

The basic formula for calculating the spread of the epidemic takes the form:

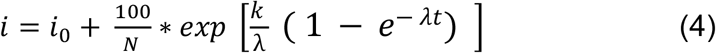

*i* - is the relative number of infected persons per one inhabitant of the settlement in question, as a percentage,

*i*_0_- is the value of i at the initial moment of the calculation period,

k. - is the transmission rate coefficient for the settlement with a population of N, which is calculated by the formula [7]:

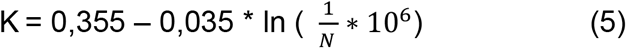

The K coefficient also depends on the transmissibility of the virus strain responsible for the spread of the epidemic during the time period in question. If the spread of infection is associated with several virus strains, the calculated relationship will be written as follows:

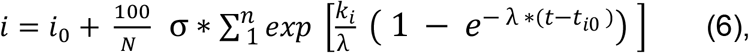

where:

i - sequence number of the strain of virus affecting the intensity of the epidemic over time *t* − *t*_*i*−*1*_,

*k*_*i*_ - transmission rate coefficient of the new virus strain and the time of the epidemic wave associated with the new coronavirus strain

*t*_*i*0_- start time of the new epidemic wave associated with the new coronavirus strain.

σ - Heaviside symbol σ = 1 When t ≥ *t*_*i*_ and σ = 0 when t < *t*_*i*_

Dependence (6) is obtained under the assumption that the two or more virus species exist independently of each other. This assumption may not hold for certain strains of the virus, in which case the calculation is performed as a sequential replacement of one virus species by another.

Numerous examples, partly presented in [4], [5], [6] and [7], show good agreement between the results of calculations using the proposed relationships (4) and (6) and statistical observations. The calculations were performed for sites as diverse as the United States, the United Kingdom or Germany, as well as for cities such as Berlin and even its individual city districts.

Correlation coefficients between the calculated and statistical data ranged from 0.94 to 0.99, confirming that the proposed calculation methodology can be used to analyse the spread of the epidemic.

However, the proposed dependencies do not take into account the impact of mass vaccination on the rate of infection. To be able to account for this crucial influence on the growth of the epidemic, the original differential equations should be slightly modified to take the following form:

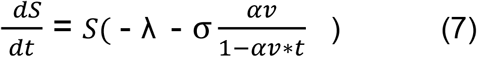

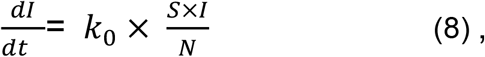

Where in equation (7) additionally:

v - population vaccination rate (1/day)

α - coefficient of vaccine efficacy,

σ - Heaviside symbol σ = 1 When t ≥ *t*_*v*_ and σ = 0 When t < *t*_*v*_,

*t*_*v*_ - is the start time of vaccination of the population

Equation (8) is formally the same as equation (2), but in reality the solution would look different because the value of S entering this equation changes. Equation (7), establishes the change in the number of persons potentially susceptible to the virus under conditions of mass vaccination of the population. The denominator in the last summand of equation (7) takes into account that as the proportion of the vaccinated population αv*t increases, the degree of impact of vaccination on the reduction of the epidemic increases. The coefficient of effectiveness α depends on both the type of vaccine and the vaccination dose (first or second). We will assume that the maximum percentage of the vaccinated population will not exceed (αv*t)max ≤0.8, i.e. with 80% vaccination of the population, an epidemic cannot develop due to the fact that some 10% of the Israeli population have already been cured of COVID 19 and over 10% have been treated asymptotically.

The solution to equation (7) is as follows:

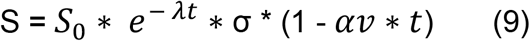

After substituting (9) into (8), solving the resulting equation, transformations and moving to a relative number of infections, we obtain the basic calculation equation for t ≥ *t*_*v*_ :

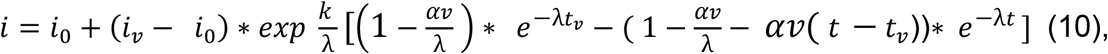

where *i* = *i*_*v*_ at t = *t*_*v*_

The calculation is carried out first by (4) or (6) and then, for the time period during which the vaccination takes place, by (10).

## Results

Let us test the proposed methodology by calculating the development and successful suppression of an epidemic by mass vaccination in Israel. By the time this work was written (mid-April 2021), about 60% of the population had been vaccinated with the first dose of vaccine, and about 55% of the Israeli population had been fully vaccinated [8].

The active phase of a coronavirus epidemic until mass vaccination begins (first half of January 2021) is calculated using equations (4), (5), and (6). With an Israeli population of about 8.79 million [9], the K-factor calculated using (5) is K = 0.43 1/day. Formula (5), which was used to determine that coefficient, was obtained for the conditions of the autumn-winter season in Europe. Climatic conditions in Israel differ considerably from those in Europe, which, according to the results of numerous studies, can have a marked effect on the growth rate of the epidemic. The most notable changes in the growth rate of COVID19 are related to the effects of the UV index and air temperature. For example, in [10], an increase in UVI decreases the maximum infection rate by about 6% per unit increase in the index. Other studies have cited a wide range of values for the effect of temperature. In [11], based on an analysis of the initial stage of an epidemic in more than 100 countries and assuming exponential epidemic development, the effect of changes in air temperature and UVI on the growth of the infection was analysed. Statistical equations for the relationship between air temperature and UVI and the growth rate of the epidemic are presented. Having made control calculations of these relationships, we found, in particular, that at temperatures of 10 degrees the epidemic growth rate reduced by 5% with an increase in temperature by 1 degree, and at temperatures around 25-30 degrees this decline is about 2%. The work also shows that the decrease in the epidemic intensity with an increase by 1 unit in the UVI value (our estimate) is about 5-7%. However, it should be noted that there is a very high correlation between these parameters, which makes it difficult to identify each of these climatic characteristics. We consider the introduction of a UVI value when adjusting the intensity of infection growth to be important, as radiation not only kills viruses, but also contributes to the population’s resistance to infection by increasing vitamin D levels [12].

In the absence of any reliable information, we take the mildest option of a 1% decrease in Δ*i*_*max*_ with an increase in temperature by 1 degree. Thus, as a first approximation, we find an estimated dependence of the value of maximum infection on climatic characteristics in the form:

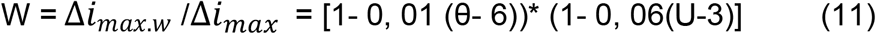

where W is the coefficient of influence of climatic parameters on intensity of epidemic development, Δ_*imax.w*_ is the maximum intensity of infection growth taking into account climatic factors, θ is average air temperature (C^0^), U is the value of UV index (for average Berlin conditions it is assumed that θ = 6^0^ C, U=3). The value of maximum growth of infection is established from dependence (4) and according to [5] has the form:

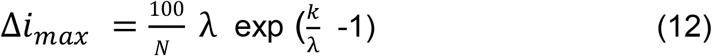

From (12) after transformations we find

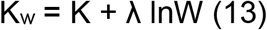

where K_w_ is the coefficient in (4), (5), (6) and (10) taking into account the influence of climatic factors. For Israel, statistical data on climatic characteristics are given in [13].

For period from April till October this factor decreases to Kw = 0.41 1/day, from November till March climate differs comparatively little from average European climate and that’s why K_w_ is taken without correction for climatic factors

Fig. 1 shows calculations using dependencies (6) and (10) and statistical data for Israel for the period 08.05.2020 to 09.04.2021.

**Fig. 1:**
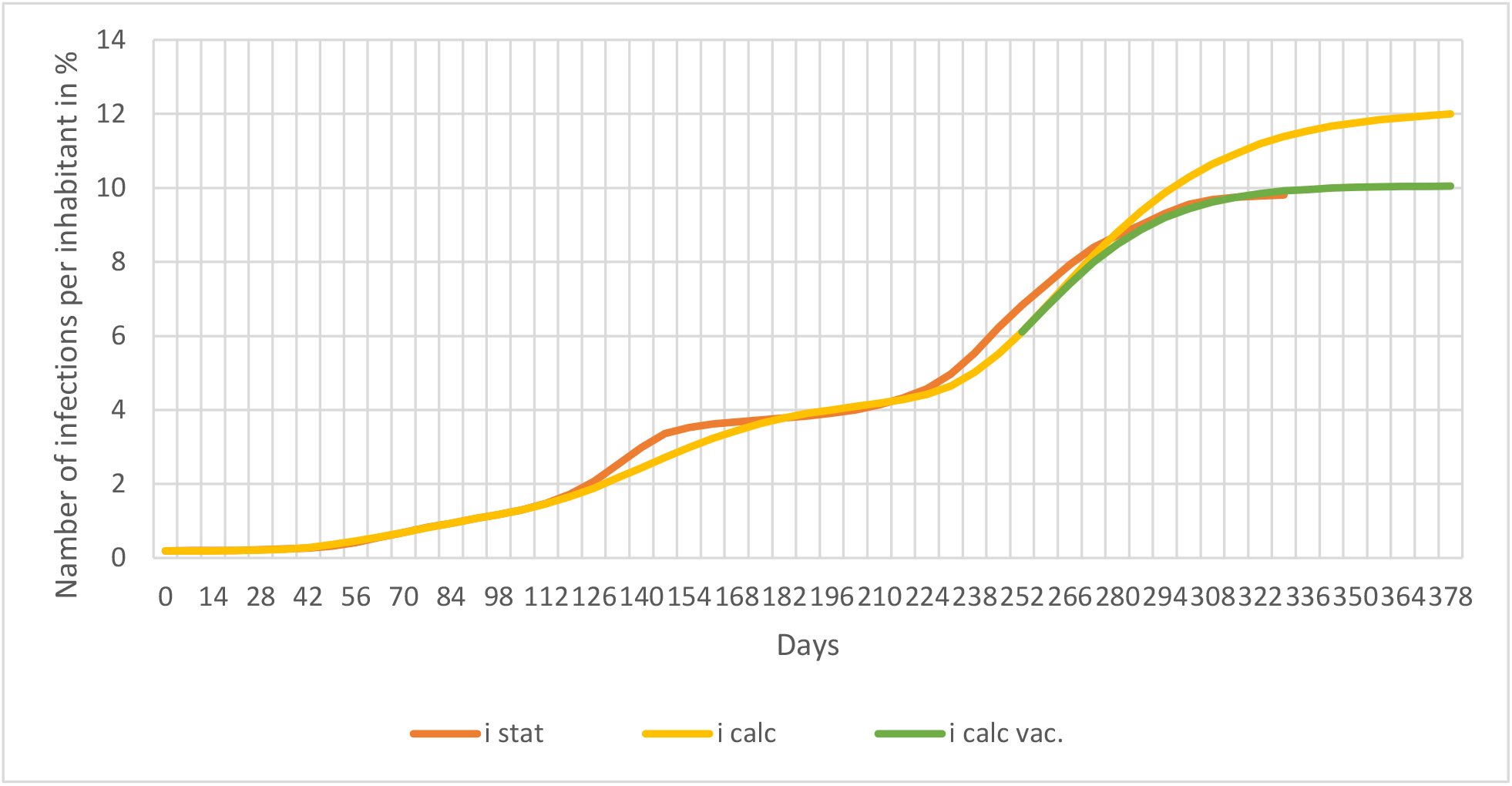
Development of the epidemic in Israel

In performing the calculations, the coefficient that takes into account the severity of lockdown for Israel was assumed to be the same as for most European countries [7] λ = 0.035 1/day.

In this figure:

i stat. - observed data,

i calc. - calculation without vaccination,

i calc. vac. - calculation for conditions of mass vaccination of the population.

The effective vaccination rate parameter was defined as the sum of the effects of both vaccine doses:

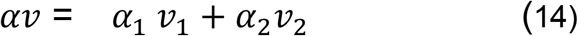

Vaccination rates for each vaccine dose were calculated from the data given in [9] as the ratio of the percentage of the population vaccinated to the total time of mass vaccination of the population. Pfizer and Moderna vaccine efficacy ratios for the first and full vaccination doses were taken as α_1_= 0.46, α_2_= 0.92 respectively [14].

For Israeli conditions, the effective vaccination rate parameter reaches a value of αv = 0, 008 1/day (for comparison: for Berlin for the same period αv = 0, 001 1/day).

The spread of the epidemic was calculated taking into account the fact that infection was associated with three major virus strains. For the first virus strain (from 08.05.2020) the calculation factor was taken as K_w_ = 0.41 1/d, for the second strain (from 24.07.2020) K_w_ = 0.435 1/d and for the third (from 20.11.2020) K_w_ = 0.47 1/d.. Accordingly, three “waves” of epidemic spread are traced. Such a high K_w_ value for the third wave of the epidemic is mainly due to the high transmissibility of the English virus strain [15] responsible for the third wave of the epidemic.

## Discussion

In general, the calculated data correspond quite satisfactorily with the statistics for the whole period of the epidemic in Israel. The correlation coefficient between estimated and observed data is higher than 0.995.

The observed number of infected persons was significantly higher than the calculated data between 112 and 140 days from the beginning of the epidemic, probably due to the influence of Rosh Hashanah in early September. In [7], a correlation is given to determine the effect of an abrupt change in lockdown conditions on the intensity of transmission. The use of that relationship would have allowed such changes in lockdown conditions to be taken into account, particularly during major holidays, but we did not consider it necessary to make such clarifications, because the main objective of the present work is to analyze the effect of vaccination on the development of an epidemic. For that purpose, model calculations were made of the relationship between i_max_ and vaccination rate. The value of i_max_ was calculated using dependence (10), assuming that *t* = *t*_*max*_. The maximum vaccination time was estimated based on the condition that no further spread of the epidemic is possible with a vaccination rate of around 80%, i.e. α*vt*_*max*_ = 0, 8 therefore *t*_*max*_ = 0, 8/ (α*v*).

The equation for calculating the i_max_ after a simple transformation is:

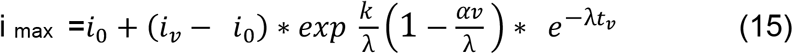

The notations in (15) are given above for relation (10).

The results of the calculations for this relationship are shown in Fig. 2.

**Fig. 2:**
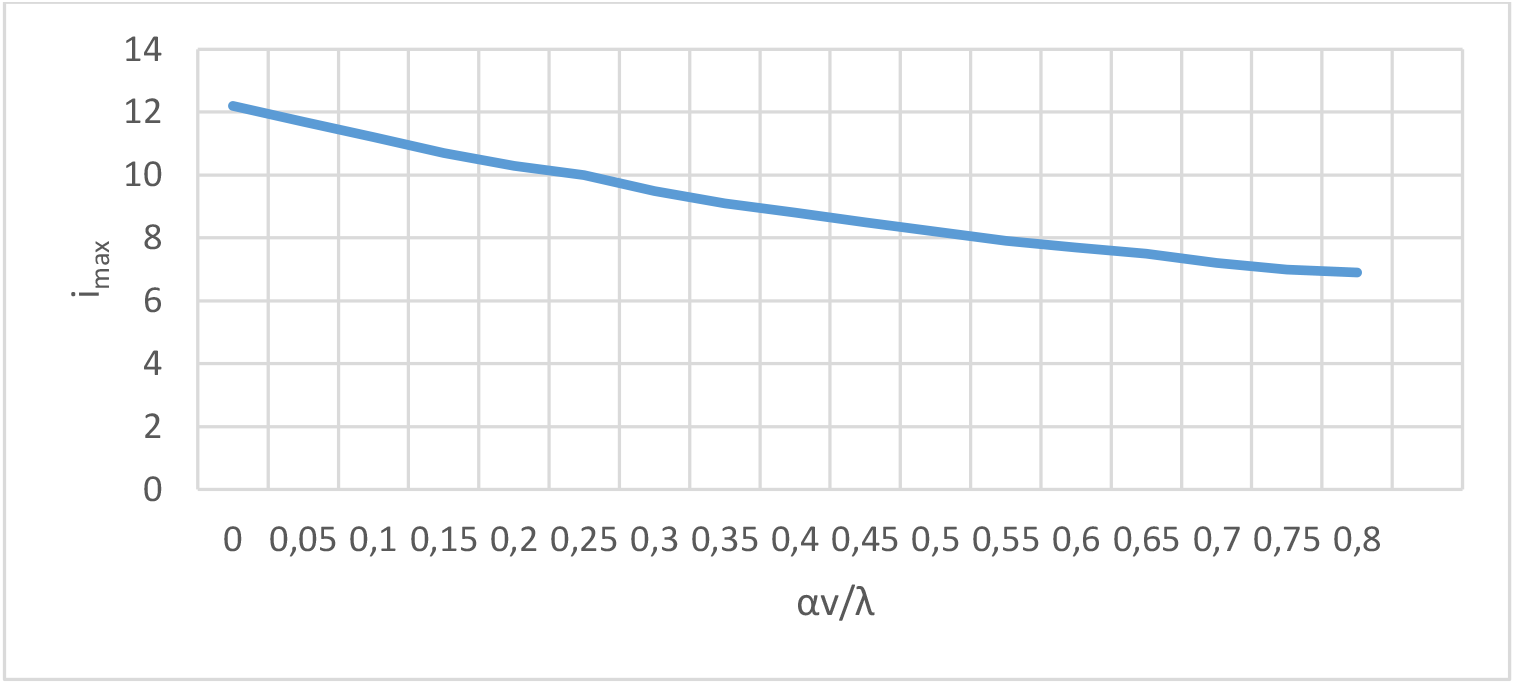
Effect of vaccination rate on the intensity of epidemic growth

The effective vaccination rate is related to the coefficient λ, which was assumed constant in the calculations and equal to λ = 0.035 1/day. As would be expected, the maximum number of infections decreases as the vaccination rate increases.

The proposed computational model for infection reduction due to vaccination yielded computational data in good agreement with observations. Mass vaccination is the most effective means of controlling the spread of virus infection. At the same time, however, even during mass vaccination a dramatic weakening of the lockdown conditions can lead to an increase in the intensity of the spread of the epidemic.

Figure 3 compares the statistics for the vaccination period with the results of calculations under different lockdown conditions.

**Fig. 3:**
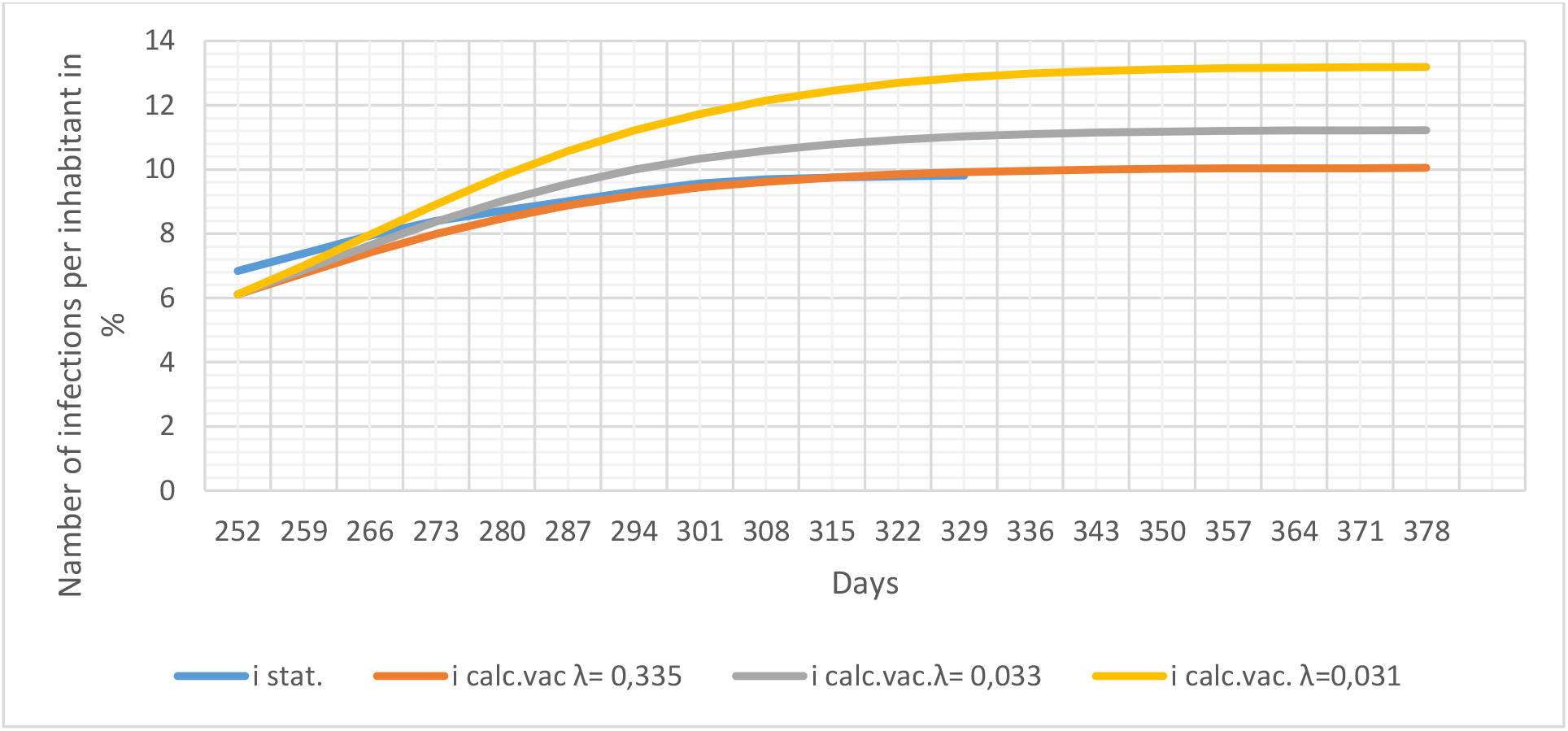
Epidemic development during the vaccination period under different lockdown conditions

The calculations were performed for the condition that lockdown is cancelled simultaneously from the start of mass vaccination, partially (λ = 0.033 1/day) or completely (λ = 0.031 1/day). (The relationship between lockdown conditions and the model coefficient λ is discussed in [7]). In both cases, an increase in the number of infections can be expected despite continuous vaccination. Moreover, in the case of complete elimination of lockdown, the model predictions suggest that the maximum number of infections would even exceed that for conditions of no vaccination but strict maintenance of lockdown (see Figure 1).

However, it should be borne in mind that when mass vaccination is carried out, the likelihood of a new strain of virus emerging and therefore a new epidemic wave is significantly lower than when the lockdown rules are followed.

An estimate of the probability of the number of mutations without vaccination of the population can be made by assuming that a significant change in the properties of the strain is a rare event. If on average 3 such mutations occur per year (our model results show that in many localities and countries 4 such mutations occur per year), using the Poisson distribution, we obtain for the probability density of new strains of the virus:

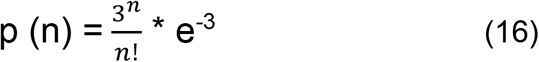

As a result of the estimation of (16), we obtain in particular that without vaccination, the probability of more than two new virus strains a year influencing the growth of the epidemic is around 60%.

We also modelled the change in the maximum number of infected persons when the lockdown is delayed by (t_L_.–t_v_.) days, instead of being mitigated or cancelled when vaccination starts. The results of these calculations are shown in Figure 4. In this figure the dimensionless time parameter T = (t_L_–t_v_) / (t_max_. – t_v_),) is given on the abscissa axis, the dimensionless complex I = (i_max_…_L_ - i_max_…) / i_max_. * 100 %, showing the magnitude of the change in the maximum infection values when the lockdown requirements are reduced. The calculation was carried out according to (15), but instead of t_v_ the time of change in lockdown conditions t_L_. was substituted in this equation. The values of the number of infected persons i_0_ and I_L_ for selected times t_L_ at λ = 0.035 1/day were calculated using equation (10).

**Fig. 4:**
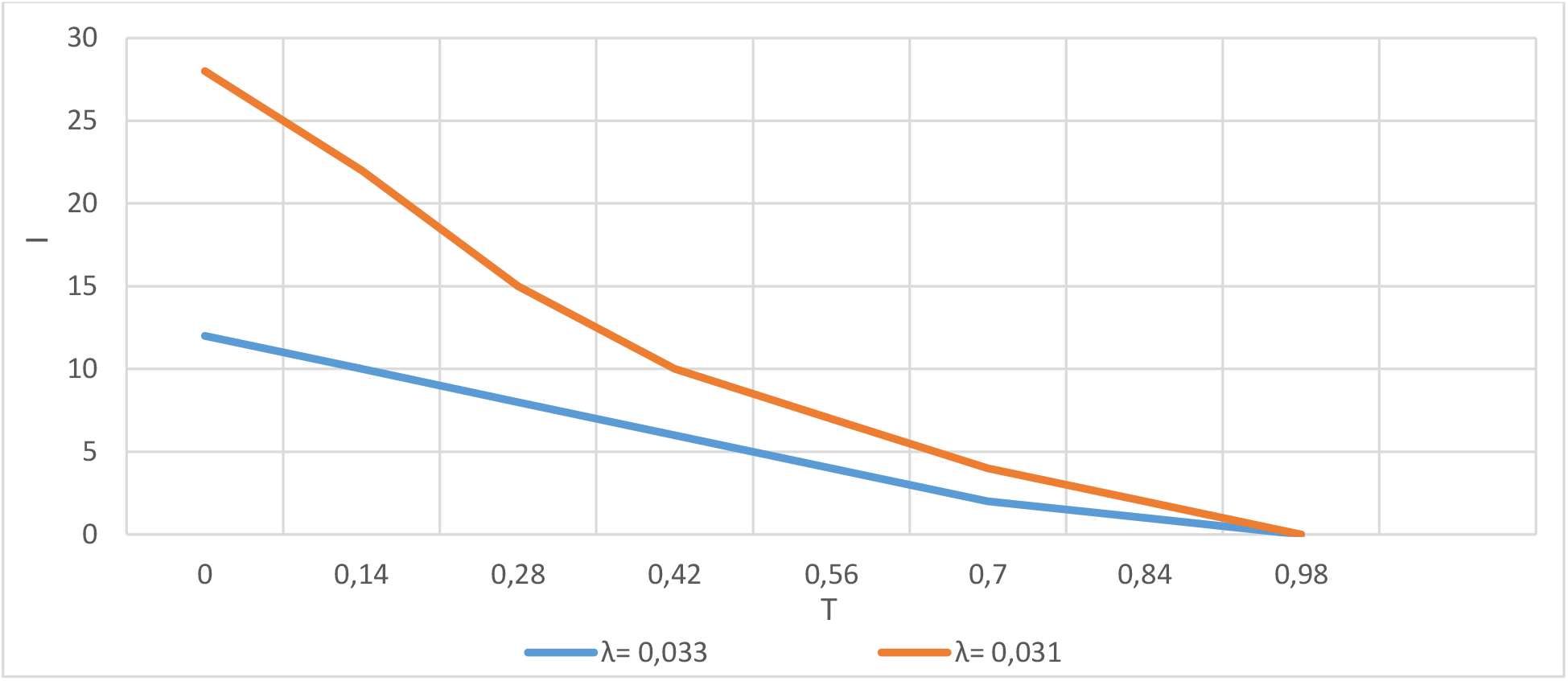
Epidemic development at different times of changing lockdown conditions during mass vaccination

When the timing of the lockdown change and the start of vaccination coincide, i.e. when T = 0, the increase in the epidemic is maximum, e.g. if the lockdown is completely cancelled, the increase in the number of infections is almost 30%. If lockdown is cancelled one month after vaccination has begun, the maximum increase in infections does not exceed 15%, and after two months, it is only about 5%. These results are obtained for the real vaccination rate in Israel. Thus, based on these calculations, it is possible to estimate the consequences of changing the conditions of lockdown compliance depending on the vaccination rate and the timing of these changes.

The epidemic process, according to the model developed, is defined by three dimensionless parameters: 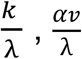 and λt. The variables that make up these complexes all characterize the magnitudes of the intensities of individual processes: K for infection, λ for reduction of infections as a result of lockdown and αv for vaccination. Each of these processes depends on many factors, some of them can be regulated, and others cannot. The K parameter, which depends on the virus strain, population size and climatic conditions, cannot be controlled. The other two parameters depend on the effectiveness of lockdown and vaccination, i.e. can and should be administratively regulated.

On the basis of these data, as well as the data presented in previous studies, it is clear that, although the model was originally developed for the initial stages of an epidemic, its high efficiency is still valid for analysing subsequent stages of infection. This important conclusion provides grounds for further research into the development of a promising predictive model for epidemic analysis. On the basis of this model, an operational as well as long-term management system can be developed, aimed at minimizing the infection rate of the population.

To further improve and refine the model, it is advisable to use it for individual age or socio-demographic groups of the population. Benchmark calculations performed for Berlin have shown that no modification of the model is required to account for age groups. The optimal vaccination plan for different age groups can be justified on the basis of epidemic growth analysis. The influence of demographic characteristics has been analysed in [4]. In particular, it was found that a higher percentage of Berlin districts with roots in countries of the Islamic Commonwealth increased the intensity of the epidemic compared to other districts. Moreover, the intensity of vaccination is also related to demographic characteristics of the population. For example, according to [16] in the United Kingdom “the lowest vaccination rates were observed among people identifying as Black African (58.8%), Black Caribbean (68.7%), Bangladeshi (72.7%) and Pakistani (74.0 %). The vaccination rate among people from an Indian background was lower than that of the White British group but remains high at 86.2%. The rate of vaccination depends not only on the availability of sufficient vaccines but also on the willingness of citizens to be vaccinated, which is related both to the level of public awareness and to the psychological state of people. Psychological factors should also be taken into account when further refining the methods used to predict the growth of the epidemic.

For the proposed model to be able to forecast the spread of infection more reliably in the long term, the parameter characterising the intensity of epidemic growth and the probability of mutation should be linked to the characteristics of the virus strains. For this purpose, the intensity of the epidemic and the sequencing of virus strains under different conditions should be investigated in parallel.

At the same time, in the form already proposed, the model developed could be an additional important tool for analysing, controlling and short-term forecasting of epidemics, depending on administrative decisions to reduce infection rates.

## Conclusions

1. A modified model of the epidemic under conditions of mass vaccination was developed. A comparison of the model results with statistical observations in Israel shows good agreement.
2. We show that the intensity of coronavirus epidemics depends on climatic characteristics, in particular air temperature and UV index. A relationship is given to estimate the impact of these factors on infection growth.
3. Three dimensionless complexes composed of transmission, vaccination and contact restriction intensities are found to determine the epidemic development.
4. Model simulations of the efficacy of limiting the epidemic by lockdown and vaccination are performed. Mass vaccination of the population is the most radical method for limiting the growth of the epidemic. Lockdown cannot completely prevent the expansion of the epidemic.
5. The probability of occurrence of new virus strains without vaccination is estimated; the probability of more than two new virus strains a year influencing the growth of the epidemic is about 60%.
6. A controlled calculation of the effect of timing of changes in lockdown conditions during the period of vaccination on the development of the epidemic. It was shown, in particular, that the cancellation of lockdown simultaneously with the start of vaccination did not reduce the maximum number of virus infections.
7. Calculations have been made on the effects of gradual lockdown cancellation. Based on these calculations, it is possible to estimate how the epidemic develops under different variants of partial lockdown cancellation.
8. The way forward for further development of the model is outlined. The immediate goal of modifying the model is to use it for each age group in the population and to find out the relationship between vaccination rate and the psychological state of the population, i.e. people’s readiness for mass vaccination.

## Data Availability

[1] Brauer, F., Castillo-Chavez, C., & Feng, Z. (2019). Mathematical models in epidemiology (Vol. 32). New York: Springer.

[2] Groendyke, C., & Combs, A. (2020). Modifying the network-based stochastic SEIR model to account for quarantine. arXiv preprint arXiv:2008.01202.

[3] Below, D., & Mairanowski, F. (2020). Prediction of the coronavirus epidemic prevalence in quarantine conditions based on an approximate calculation model. medRxiv.

[4] Below, D., Mairanowski, J., & Mairanowski, F. (2020). Checking the calculation model for the coronavirus epidemic in Berlin. The first steps towards predicting the spread of the epidemic. medRxiv.

[5] Below, D., Mairanowski, J., & Mairanowski, F. (2021). Analysis of the intensity of the COVID-19 epidemic in Berlin. Torwards an universal prognostic relationship. medRxiv.

[6] Below, D., Mairanowski, J., Mairanowski F. (2021) Comparative analysis of the spread of the COVID 19 epidemic in Berlin and New York City based on a computational model. https://www.medwinpublishers.com/PHOA

[7] Below, D., Mairanowski, J., Mairanowski F. (2021). Development of the COVID-19 epidemic model: calculations for a mutating virus. https://medwinpublishers.com/JQHE/

[8] Statistics and Research: Coronavirus (COVID-19) Vaccinations (2021). https://ourworldindata.org/covid-vaccinations

[9] List of countries and dependent territories of the World by population (2021). https://countrymeters.info/en/list/List_of_countries_and_dependent_territories_of_the_World_by_population

[10] Moozhipurath, R. K. (2020). Role of Weather Factors in COVID-19 Death Growth Rates in Tropical Climate: A Data-Driven Study Focused on Brazil. medRxiv.

[11] Notari, A., & Torrieri, G. (2020). COVID-19 transmission risk factors. arXiv preprint arXiv:2005.03651.

[12] Ilie, P. C., Stefanescu, S., & Smith, L. (2020). The role of vitamin D in the prevention of coronavirus disease 2019 infection and mortality. Aging clinical and experimental research, 32(7), 1195-1198.

[13] Weather Atlas Resources (2021). https://www.weather-atlas.com/en/resources.

[14] Dagan, N., Barda, N., Kepten, E., Miron, O., Perchik, S., Katz, M. A., … & Balicer, R. D. (2021). BNT162b2 mRNA Covid-19 vaccine in a nationwide mass vaccination setting. New England Journal of Medicine, 384(15), 1412-1423.

[15] Davies, N. G., Abbott, S., Barnard, R. C., Jarvis, C. I., Kucharski, A. J., Munday, J. D., … & Edmunds, W. J. (2021). Estimated transmissibility and impact of SARSCoV-2 lineage B. 1.1. 7 in England. Science, 372(6538).

[16] Vahé Nafilyan, Charlotte Gaughan and Jasper Morgan (2021). Coronavirus and vaccination rates in people aged 70 years and over by socio-demographic characteristic, England: 8 December 2020 to 11 March 2021. Office for National Statistics.

## Notes

### Competing Interest Statement

The authors have declared no competing interest.

### Funding Statement

No external funding was received.

### Author Declarations

All relevant ethical guidelines have been followed.

